# “Addressing the core trait of bipolar disorder”: a concept analysis of mood stabilizing drugs

**DOI:** 10.1101/2021.11.13.21266303

**Authors:** Lauro Estivalete Marchionatti, Paula Blaya-Rocha, Pedro Vieira da Silva Magalhães

## Abstract

**Background:** The term “mood stabilizer” is controversial in the literature. As there is no consensual meaning, its retirement has been suggested to avoid misuse. Nevertheless, it remains largely employed, and may carry an important meaning. This issue has not been approached using a validated qualitative inquiry.

**Methods:** We employed document analysis for reviewing definitions for mood stabilizer. Then, we used concept analysis as a qualitative methodology to clarify the meanings associated with the term. Based on its results, we built a theoretical model for a mood stabilizer, matching it with evidence for drugs used in the treatment of bipolar disorder.

**Results:** Concept analysis of documents defining the term unearthed four attributes of a mood stabilizer that were nested into the following ascending hierarchy: “not worsening”, “acute effects”, “prophylactic effects”, and “advanced effects”. To be considered a mood stabilizer, a drug had to reach the “prophylactic effects” tier, as this was discussed by authors as the core aspect of the class. After arranging drugs according to this scheme, “lithium” and “quetiapine” received the label, but only the former fulfilled all four attributes, as evidence indicates it has neuroprotective action.

**Conclusion:** The proposed model uses a hierarchy of attributes that take into account the complexity of the term and help to determine whether a drug is a mood stabilizer. Prophylaxis is pivotal to the concept, whose utility lies in implying a drug able to truly treat bipolar disorder, as opposed to merely targeting symptoms. This could modify long-term outcomes and illness trajectory.

## INTRODUCTION

The moniker “mood stabilizer” appeared sparsely and occasionally referring to lithium until the late 80s (Kerry and Owen 1970). While Schou (1963) had proposed that mood normalizers would “turn a pathologically changed mood into a normal one”, the later notion of mood stabilization was more akin to prophylaxis and disease modification; stabilizing means “making unlikely to fail”, a stabilizer “a thing used to make something steady or stable”. With the incorporation of carbamazepine and valproate into the arsenal used to treat bipolar disorder and the new focus on prophylaxis, the “mood stabilizers” became a new class of psychotropics. In the 90s, the label gained popularity, being extended to antipsychotics (Zarate et al. 1995; Davis John M and Janicak Philip G 1996), with suggestions it was mainly chosen pragmatically by the pharmaceutical industry to imply new drugs were as effective as lithium (Healy 2008, chap. 6).

More recently, the utility of the term has been repeatedly questioned, as one that lacks clarity and consequently is loosely employed (Safer 2010; Ghaemi 2001; G. M. Goodwin and Malhi 2007). The peril here would be that medications be labeled mood stabilizers after showing efficacy in specific domains of bipolar disorder and, once thus branded, their use extended beyond the clinical domain its efficacy was demonstrated (Malhi et al. 2018; Malhi and Roy Chengappa 2017). This has been used as a further argument for retiring the term (Malhi and Roy Chengappa 2017; Safer 2010).

An analysis of trends of its use in titles of scientific papers does suggest a decline. From 1987 (its first recorded use in a title of a scientific paper), it has increased greatly until 2002, to then stabilize thereafter, a significantly different trajectory to the use of “bipolar disorder” (See Figure 1 and Figure 2 for comparison). Major professional guidelines still tend to employ the term liberally, although often disclaiming that the term ought to be employed more carefully and clearly. To cite just one instance the latest Canadian Network for Mood and Anxiety Treatments (CANMAT) guidelines state that its “use in the literature is inconsistent, and so [it] will not be used” (Yatham et al. 2018). Nevertheless, the term then appears another 22 times throughout the document, and this is analogous to several other professional guidelines (Malhi, Outhred, et al. 2018; National Collaborating Centre for Mental Health (UK) 2018). This may suggest that the term “mood stabilizer” cannot be easily set aside, as it may still communicate some needed meaning. Perhaps, as Malhi et al. (2018) posit, it offers some comfort to patients and clinicians. It indeed appears on many patient education pages, such as those of the National Health Service (14 March 2019), National Institute for Health (2016), and UptoDate (Stovall 2021), to name a few.

**Figure 1.**
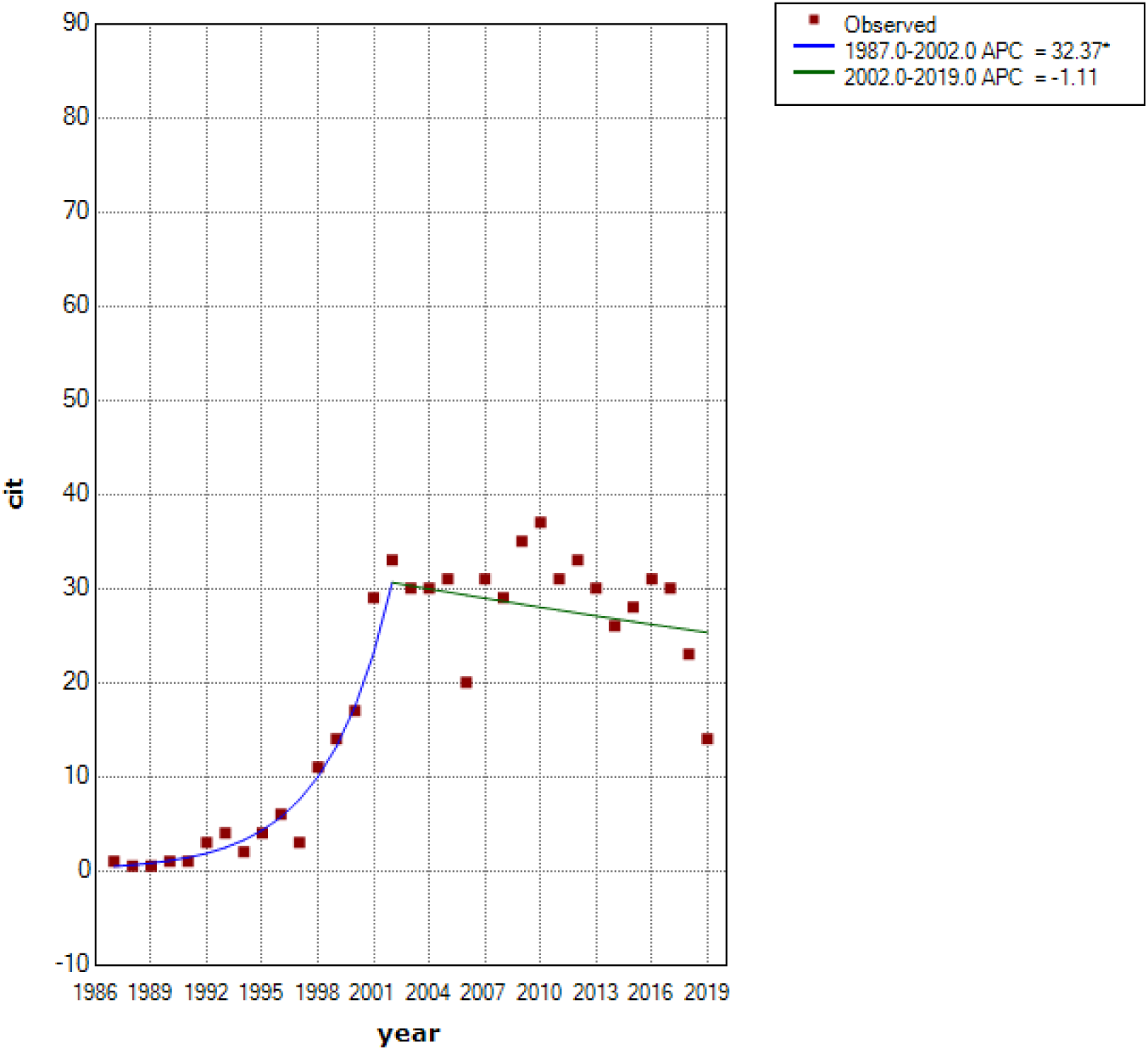
The number of publications per year including the term “mood stabilizer” in Medline database. There is one joinpoint indicating that the Annual Percent Change is significantly different from zero at the alpha = 0.05 level.

**Figure 2.**
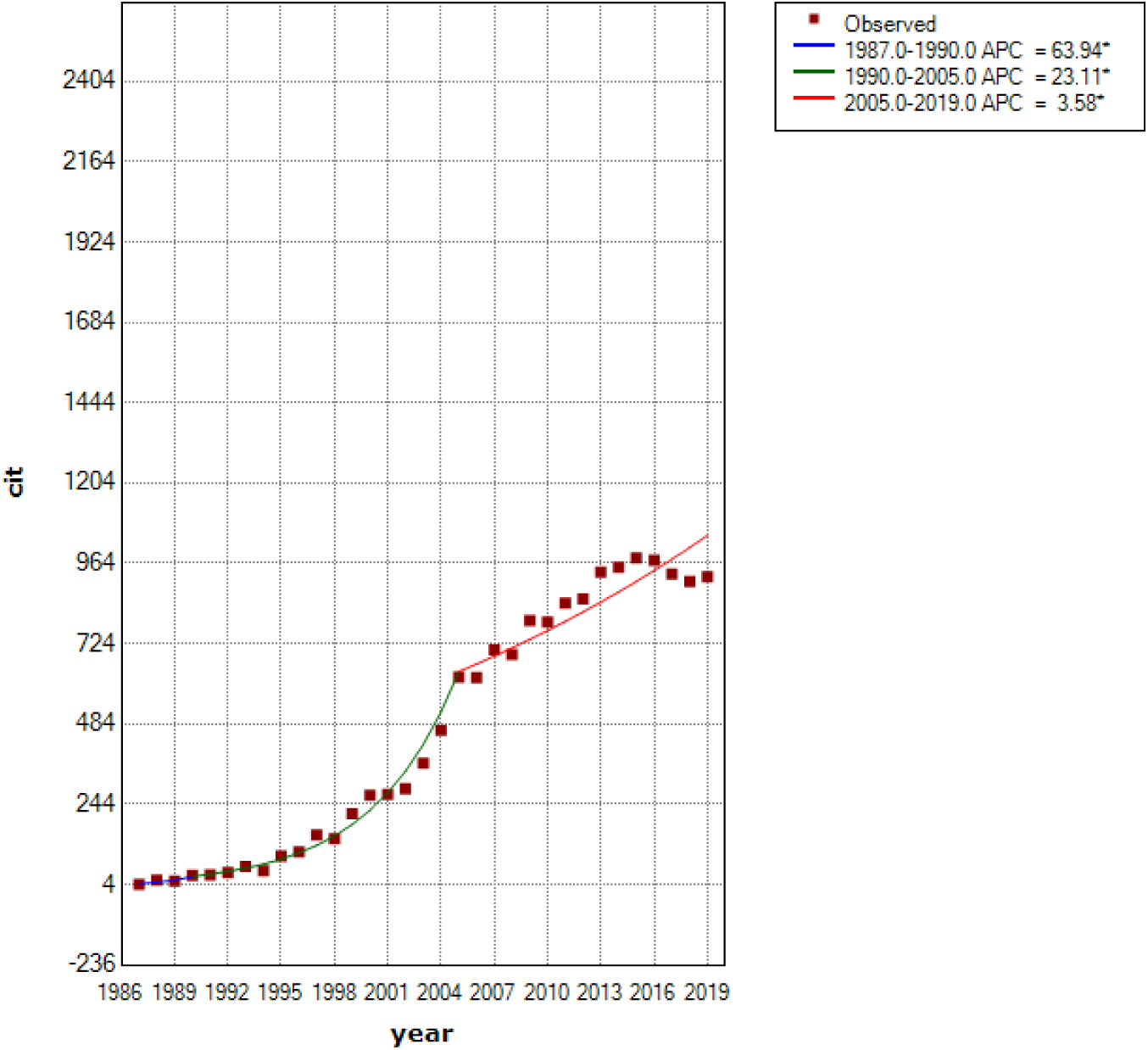
The number of publications per year including the term “bipolar disorder” in Medline database. There are two joinpoints indicating that the Annual Percent Change is significantly different from zero at the alpha = 0.05 level.

The utility and destiny of this “class” of medications have certainly been long debated, most recently by Malhi and Chengappa (2017) in a series of thoughtful comments. While we see merit in arguments from both sides of the aisle, some conceptual work could help in pointing to a more precise definition of what authors propose mood stabilizers should be. Here, we sought to define more clearly the meaning of the “mood stabilizer” epithet and the consequences of this classification. We began by using document and concept analysis to delineate the necessary attributes of the term. While the usage of the term has been examined before, we are unaware of a formal exploration such as the one we provide here. Next, we arrange and propose a tentative hierarchy of properties based on the categories we found. Finally, we made an attempt to operationalize these categories and, using the best available evidence, see where currently available treatments for bipolar disorder stand in this hierarchy.

## METHODS

We employed document analysis as described by Bowen (2009) as the procedure for reviewing the material. The procedure entails finding, selecting, evaluating, and synthesizing the data contained in documents; the passages are then organized into themes and categories. We looked specifically for conceptual definitions through skimming, reading, and discussing for the planned concept analysis. To be included, a document should specifically address and propose a clear definition of a “mood stabilizer”. Thus, inclusions were mostly narrative reviews in which the authors could have used any number of rationales to come up with a definition of a mood stabilizer. The documents of interest were mostly published articles, but we also included books addressing the issue when referenced. We undertook searches in Medline, Scopus, PsychInfo, and Google Scholar with the terms “mood stabiliz*”, “mood stabilis*”, “mood normaliz*”, without constraints on time of publication or language. While admittedly not systematically reviewing everything written on mood stabilizers, we sought to include recognized proposals and reference guidelines and inspected reference lists on those for additional definitions. As such, we excluded from the concept analyses those pieces that only review or criticize the term (for instances, see Sobo (1999), Keck and McElroy (2003), and López-Muñoz et al. (2018)), although we address some of these in the discussion. Document analysis does not rely on completeness, but on a wide array of documents, prioritizing quality over comprehensiveness.

### Concept analysis

This is a strategy used for examining the semantic structure of concepts, determining attributes and characteristics and then arriving at precise theoretical and operational definitions (Foley and Davis 2017). By delineating characteristics of a concept, the researcher can formulate precise theoretical definitions to be used in further studies, serving as a firmer starting point to guide research and exploration.

There are several approaches to conducting a concept analysis (Walker 2005; Morse 1995; Wilson 1963). We employ evolutionary concept analysis as described by Rodgers and Knalf (1993), which presents a clear analytic framework, with strengths in its systematization and with well-established steps. It takes into account how a concept evolves over time, considering it a dynamic process wherein the context is central. Briefly, the analysis consists of identification of a concept of interest including surrogate terms, selection of a sample for data collection, collection of data relevant to attributes of the concept (including its context, antecedents, and consequences, as well as interdisciplinary and sociocultural variations), analysis of data according to these characteristics, identification of an exemplar, and identification of hypotheses and implications for further development of the concept. Each piece was read several times until the general theme was identified; analysis proceeded using phrases as units of analysis.

### Operationalization for efficacy analysis

The strength of the evidence is a further consideration to evaluate potential mood stabilizers (Malhi et al. 2018). Thus, as a next step, we sought to operationalize the categories obtained from the content analysis and look for the best evidence in the extant efficacy literature for drug effects in bipolar disorder, following Bauer and Mitchner (2004). We opted to describe treatment response per phase (acute vs maintenance) and polarity (manic/mixed vs depressed). We are aware, as Malhi et al. (2018) note, that other dimensions such as activation are involved as well, but this is how authors tend to conceptualize response when conducting studies. Whenever possible, we drew on network meta-analysis of monotherapy trials as the most comprehensive and simplest methodology to determine efficacy in such phases, following evidence hierarchy (Roever and Biondi-Zoccai 2016). This supplied data referring to the treatment of acute depression (Bahji et al. 2020a), acute mania (Yildiz et al. 2015), and maintenance treatment (Miura et al. 2014). However, randomized placebo-controlled studies concerning long-term data are sparse and usually restricted to relapse rates, providing limited information. In these cases, we looked further into the naturalistic hospitalization data. While many such reports understandably use lithium as a relevant comparison (Kessing et al. 2018), more pertinent to our objectives were studies that employed an on-off design, testing the hypothesis of a benefit or harm of different medications. Joas et al. (2017) tested a number of drugs in Swedish registries considering their benefits in preventing hospitalizations, while Lähteenvuo et al. (2018) included an assessment of detrimental effects using rehospitalization data from the Finnish population.

The “not worsening” category was operationalized considering both short-term and long-term outcomes. Short-term “not worsening” was informed by network meta-analyses of acute mania and bipolar depression, wherein a drug could not display a propensity to cause acute manic or depressive switches as being inferior to placebo. As for long-term data, while the network meta-analysis on maintenance treatment was taken into account, we further examined the evidence for harm of medication by looking at the observational on/off design by Lähteenvuo et al. (2018), which indicated whether adding a drug into a regimen increases odds of rehospitalization. Drugs with evidence of harm were classified as “mood destabilizers”, and therefore placed out of the hierarchy rank, regardless of possible benefits in subsequent categories.

As for “acute effects”, we used data from the most recently available network meta-analyses to investigate the superiority to placebo in monotherapy in acute mania (Yildiz et al. 2015) or bipolar depression (Bahji et al. 2020b), discarding those drugs previously shown to worsen outcomes.

To determine “prophylactic effects”, we started from data provided by network meta-analysis to show superiority in monotherapy to placebo in terms of relapses to mania, depression, or either (Miura et al. 2014). This category was also informed by available on/off hospitalization data based on national registries to gauge prophylactic effects on hospitalizations for mood episodes of each polarity, as informed by Joas et al. (2017). To qualify for the rank, a drug had to display efficacy in preventing episodes of either mania or depression.

The “advanced effects” category proved to be a much more difficult dimension to operationalize in terms of direct comparisons and generalizability. Instead of summing up the evidence, we limited ourselves to discussing this notion considering studies possibly signaling neuroprotective effects, as this is a feasible proxy to evaluate such outcomes.

## RESULTS

### Concept analysis

We located 8 relevant articles for our document analysis. The first relevant conceptual proposal was published in 1996 and the most recent in 2018 (Figure 3). We identified four non-overlapping attributes in the definition of a mood stabilizer.

**Figure 3.**
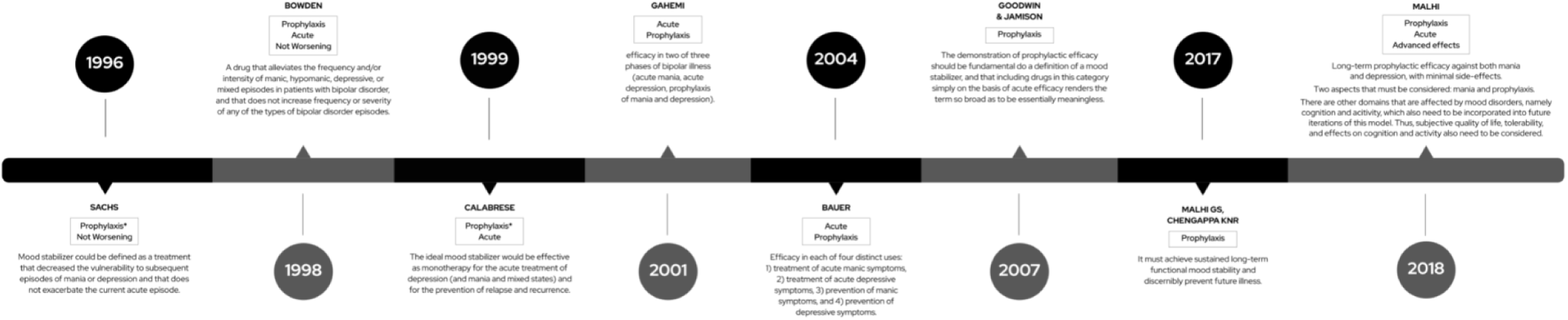
Timeline of proposed definitions.

#### Not worsening

Authors agree that mood stabilizers should not have deleterious effects on current or subsequent mood episodes. Sachs (1996) proposes that a mood stabilizer “does not exacerbate the current acute episode”, providing examples of medications that would be excluded by this rule: “available antidepressant medications may precipitate mania or worsen the course of illness […] and chronic use of antipsychotics appears to increase the risk of recurrent depression”. Bowden (1998) states that a mood stabilizer “does not increase frequency or severity of any of the types of bipolar disorder episodes”. There are clear consequences to including this category, as a drug could be beneficial in one phase, such as acute depression or mania, whilst causing affective switches or worsening long-term outcomes. Drugs that have the potential to be harmful have usually been termed “mood destabilizers”.

#### Acute effects

Authors also discuss what beneficial effects a mood stabilizer should have on the treatment of acute mood episodes. Calabrese and Rapport (1999) suggest that these drugs are “effective as monotherapy for the acute treatment of depression (and mania and mixed states)”, similarly to Bauer and Mitchner’s (2004) proposal that includes “the treatment of acute manic symptoms’’ and “the treatment of acute depressive symptoms’’. This common attribute we named “acute effects’’.

#### Prophylactic effects

Several authors discuss the properties of reducing either the occurrence, frequency, or severity of new episodes. Sachs (1996) defends that these drugs “could be defined as a treatment that decreased the vulnerability to subsequent episodes of mania or depression”, whilst Malhi and Chengappa (2017) argue “that it must achieve sustained long-term functional mood stability and discernibly prevent future illness”. We grouped these notions under a category we named **“**prophylactic effects”.

Relapse prevention is also discussed as a *sine qua non* condition for a mood stabilizer. For instance, Goodwin and Jamison (2007) state that “a prophylactic efficacy should be fundamental to the definition of a mood stabilizer, and that including drugs in this category simply on the basis of acute efficacy against depression and or mania renders the term so broad as to be essentially meaningless”. Medications with prophylactic effects are also described by Keck and McElroy (2003) as addressing “the core trait of bipolar disorder, its cyclic nature and propensity for recurrence”. They also state that “maintenance treatment of bipolar disorder is perhaps its most important aspect”. Sachs (1996) also emphasizes the primacy of prophylaxis when writing that these treatments would “result in a decreased risk of cycling whether or not they possess acute antimanic or antidepressant effects”.

#### Advanced effects

Finally, we identify a category pertaining to other effects a mood stabilizer might possess, beyond those previous categories. They were first mentioned by Keck and McElroy (2003). When summarizing uses of the term mood stabilizer, they include “comprehensive definitions” that deal with symptoms related to cognition and behavior. It was Malhi et al. (2018) who formally proposed such an attribute as part of a mood stabilizer, fleshing out that “subjective quality of life, tolerability, and effects on cognition and activity also need to be considered when determining whether a medication is a potential mood stabilizer”.

### A hierarchy of mood-stabilizing drugsã

The manner in which many authors define a mood stabilizer revealed several attributes of this class that should be taken into account when constructing a model for the concept. Individually discussing the attributes would be insufficient, as they were clearly interrelated. Some attributes were more refined and considered essential to qualify a drug as a mood stabilizer, whilst others did not guarantee the label by themselves. Therefore, we propose here a hierarchy of progressively specific properties that embraces the complexity of the treatment of bipolar disorder and could help understanding whether a drug has mood-stabilizing properties (Figure 4).

**Figure 4.**
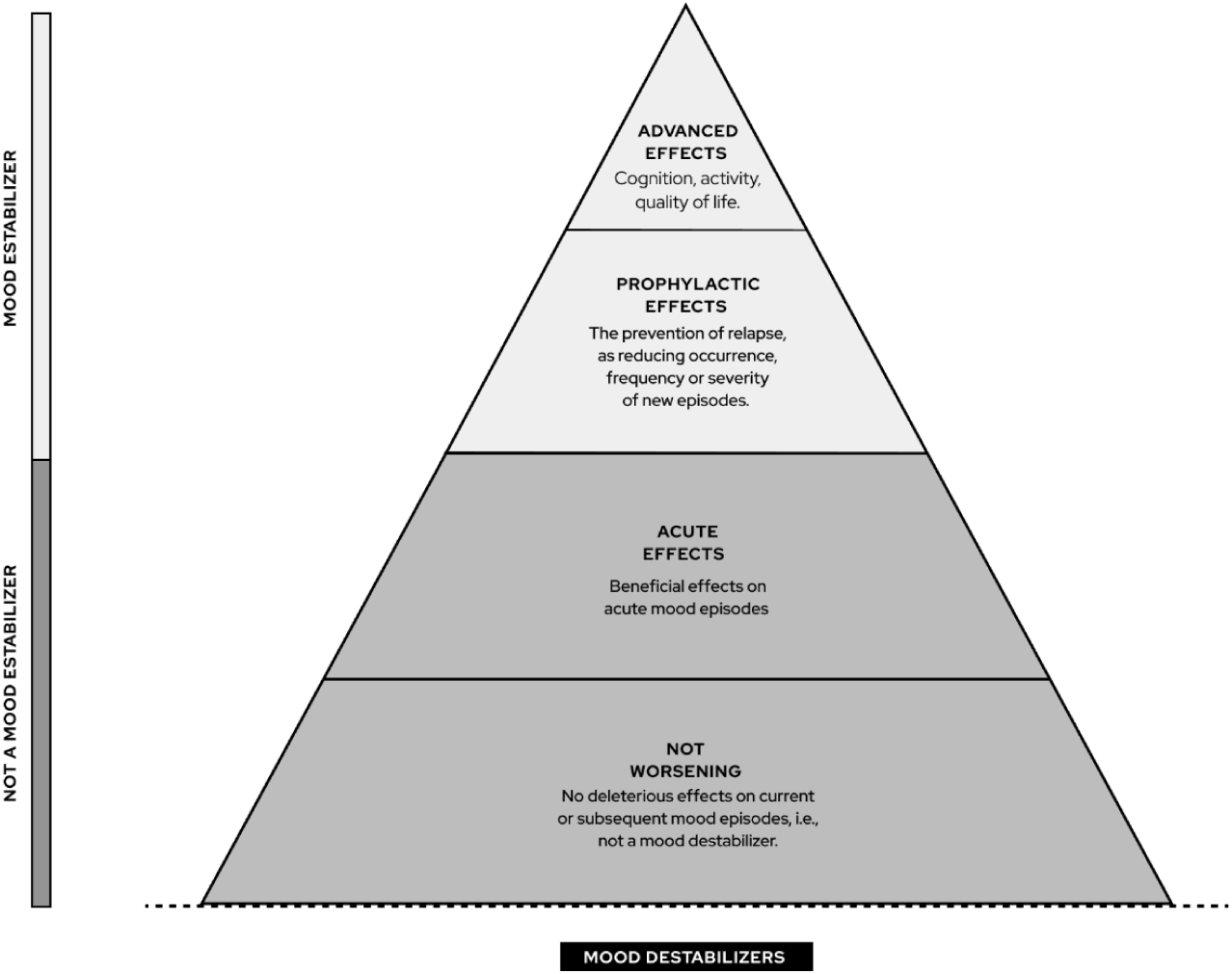
Hierarchy of a mood stabilizer: concepts.

Our first category of “not worsening any episode” seems to be cardinal to define a base: without fulfilling this precondition, a drug cannot be labeled a mood stabilizer. This tier holds a place for drugs showing neither benefit nor harm in the management of bipolar disorder. Treatments placed below this level would conceptually be outside of the hierarchy of drugs used to treat bipolar disorder. Those have usually been called “mood destabilizers”, where evidence indicates they can worsen the course of illness or cause affective switches. In the next tier, we place drugs that have no evidence of being harmful and have relevant acute effects, either for mania or bipolar depression. While not conceptualized as mood stabilizers, they could still be useful for managing acute episodes.

The next tier would consist of actual mood stabilizers, that is, drugs that would not be harmful, have acute properties for either mania or bipolar depression, and possess prophylactic efficacy for preventing mood episodes of both polarities. Finally, the apex of the pyramid would be reserved for those members that have additional desirable properties to that of a conventional mood stabilizer. These advanced effects could be understood as evidence that the drug has some fundamental ability to change the course of the illness for the better, perhaps preserving functioning and cognition.

### Current drugs used in the treatment of bipolar disorder

Finally, we gathered and organized the evidence on the effects of drugs currently used to treat bipolar disorder based on this proposed hierarchy. Figure 5 displays how drugs were allocated in our hierarchy.

**Figure 5.**
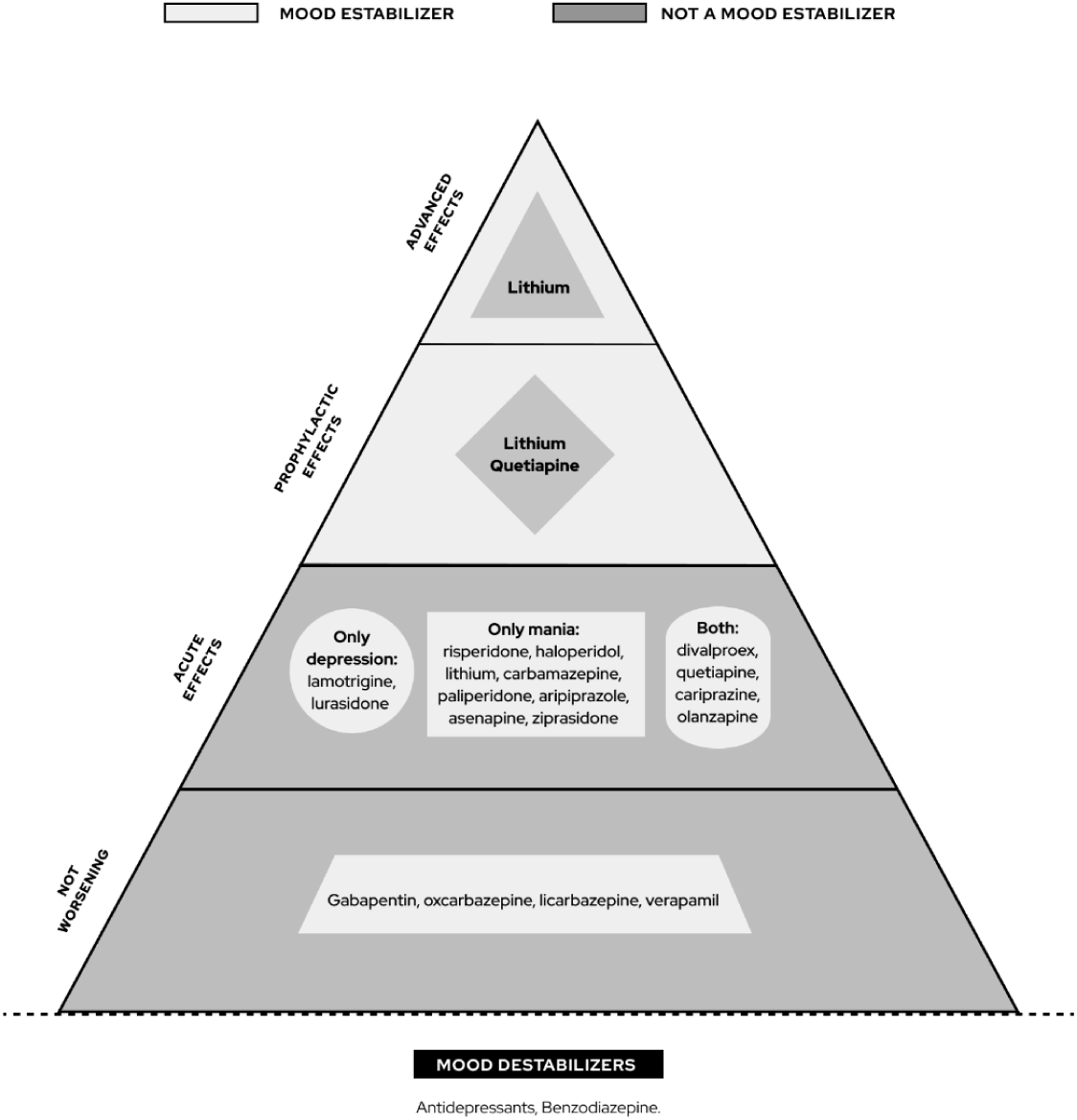
Hierarchy of a mood stabilizer: drug allocation.

Mood destabilizers should demonstrably worsen any phase or the general illness course. Network meta-analyses of maintenance treatment and acute effects did not reveal any substance significantly worse than placebo. The observational on/off literature, though, suggested hospitalization is more common during periods on any antidepressant or benzodiazepine. These drugs could be tentatively seen as mood destabilizers. The “not worsening” category was left for drugs that have been tested but did not display any evidence of harm or benefit; currently, they are gabapentin, oxcarbazepine, licarbazepine, and verapamil.

Many drugs have shown meaningful acute effects, especially for mania. These include several atypical antipsychotics, haloperidol, carbamazepine, valproate, and lithium. For bipolar depression, there are fewer drugs with randomized evidence of efficacy: four atypical antipsychotics (lurasidone, quetiapine, cariprazine, olanzapine), valproate, and lamotrigine. While some antidepressants were of benefit in treating acute depressive symptoms, they did not qualify for this rank for previously showing evidence of being mood destabilizers.

A subgroup of drugs with acute effects also showed evidence of efficacy for the prevention of episodes. Lithium, valproate, lamotrigine, quetiapine, risperidone and olanzapine were superior to placebo in preventing relapse of either mania or depression, but only lithium and quetiapine proved efficacious for preventing episodes of both polarities. The observational data by Joas et al. (2017) supported four drugs for preventing hospitalizations for both manic and depressive episodes: lithium, valproate, quetiapine, and olanzapine. As the medications that proved efficacious in both study designs and for both polarities, quetiapine and lithium qualified for the higher place of “prophylactic effects” in the hierarchy. Noteworthy, all drugs in the “prophylactic effects” category had fulfilled the previous category of “acute effects”.

As for advanced effects, we considered these properties are still not consolidated and warrant further investigation. Some studies do suggest specific effects for lithium, such as a randomized clinical trial comparing quetiapine and lithium effects on white and gray matter volume after a first episode of mania signaling possible neuroprotective effects of lithium (Berk et al. 2017). Some observational data also suggest excellent lithium responders have preserved cognitive function (Rybakowski and Suwalska 2010).

## DISCUSSION

“Mood stabilizer” is a term increasingly employed with skepticism about its utility and growing dissension as to its meaning. Our concept analysis suggested four main attributes of a mood stabilizer, which we attempted to order in a hierarchy of treatment effects: “not worsening”, “acute effects”, “prophylactic effects”, and “advanced effects”. Authors further discussed prophylactic effects as the core attribute for this class of medications. We tentatively arranged widely available drugs according to such a hierarchy using the best available evidence. Only lithium and quetiapine would fulfill the requisites to be considered mood stabilizers.

The concept of a mood stabilizer drug has been discussed before, although not using a formal document analysis such as this one. There is lingering debate on what really is a mood stabilizer (Sachs 1996; Calabrese and Rapport 1999; Bowden 1998; Ghaemi 2001; F. K. Goodwin and Jamison 2007; Malhi, Porter, et al. 2018). Regarding its historical development, Harris et al. (2003) traced its roots back to the time Schou (1963) noted lithium had prophylactic effects on manic-depressive episodes, later evolving with the emergence of anticonvulsants as first-line treatments for the disorder.

In the definitions discussed here, authors have not been satisfied in proposing a single action as the key property of a mood stabilizer. The complexity may reflect the nature of bipolar disorder psychopathology, with multiple manifestations. That is mirrored in proposals to define mood stabilizers such as the one by Malhi et al. (2018), wherein a 4×4 graph is built to quantify how effective a drug is in the prevention and acute treatment of each pole of disease, disregarding how they work in combination or what are its core properties. Nevertheless, it is precisely in dealing with this complexity that may lie the value of the concept: it implies a drug able to bring therapeutic effects for bipolar disorder, as opposed to merely targeting symptoms or specific phases of disease. Somehow related to it is the idea of “normothymotics” or “mood normalizer”, brought by Schou (1963) and “representing a class of drugs with an action specific to a disease rather than a symptom or syndrome”. This may have clear consequences for both research and clinical practice: such a drug would result in a simplified prescription and better outcomes in the long run. Indeed, we found prophylaxis to be broadly argued as a core attribute of a mood stabilizer.

Emphasizing long-term outcomes as opposed to short-term symptomatic treatment of bipolar disorder is significant in this context. Not only can mood episodes directly bring biological, psychological, and psychosocial deleterious effects to patients, but also increase the vulnerability to new episodes (Park et al. 2018). A substantial proportion of patients are thought to have a progressive illness course (van der Markt et al. 2019), and a mood stabilizer could preclude this “neuroprogression”, being a disease-modifying therapy. Helping patients achieve remission on monotherapy is another argument for keeping the concept. A medication truly treating bipolar disorder diminishes the need of employing extensive polypharmacy, each targeting some symptom or phases of the disease (Kim et al. 2021; Grover et al. 2021; Fung et al. 2019).

This debate on properties of mood stabilizing drugs is now quite a few decades old. Notwithstanding, some categories were based on the perspectives of only a few authors. Many definitions were not richly described or examined, also limiting conceptual analysis. Studies for acute effects were more readily available and provided simpler answers concerning antimanic and antidepressant effects. However, researching maintenance/prophylactic effects require long-term studies, which are more difficult and dispendious to conduct. The maintenance meta-analysis included studies from 17 weeks to 2.5 years of durations, which is shorter than expected to determine disease-modifying effects. Therefore, we supplemented it with data from on/off naturalistic studies. This method could have introduced some inherent biases of observational data, including significant interpretation challenges because of intense polypharmacy. Moreover, the advanced effects category represented a major challenge to operationalize and could not be tested with confidence. Available literature only provided clues regarding effects on cognition, mostly through an intermediate outcome of neuroprotective action. In the case of quality of life, this category could not be operationalised, as no study succeeded in establishing which drugs are associated with benefits on such outcomes.

Our concept analysis indicates the concept of mood stabilizing drugs has had multiple attributes when described by different authors over the decades. The most coherent definitions propose prophylactic effects as a fundamental property, and we believe this could potentially redeem the term, linking it to recent discussions on neuroprogression and neuroprotection. We further propose that definitions posed so far could be understood as a hierarchy of mood stabilizing properties, which could help determine whether a drug qualifies for the label. If mood stabilizers can indeed be shown to differentially affect illness course, perhaps preserving cognition and functioning, that would be the standard needed to consider specific drugs for bipolar disorder.

## Data Availability

All data produced in the present work are contained in the manuscript.

## Acknowledgments

We thank Samanta Duarte for designing the graphical representations included in this paper.

